# Parental income gradients in child and adolescent mortality: Norwegian trends over half a century

**DOI:** 10.1101/2022.03.21.22272688

**Authors:** Miriam Evensen, Søren Klitkou, Mette Tollånes, Petur B Juliusson, Øystein Kravdal

**Author notes:** **Address correspondence to:** Miriam Evensen, Institute for Social Research, PO Box 3233 Elisenberg, NO-0208, Norway.

## Abstract

**Background:** Child mortality declined rapidly the last century in many high-income countries. However, little is known about the socioeconomic differences in this decline, and whether these vary across causes of death.

**Methods:** We use register data including all Norwegian births between 1968 to 2010 (2.1 million), and analyse how all-cause and cause-specific child (0-5) and adolescent mortality (6-21) vary with relative parental income the year before the birth

**Results:** Child all-cause mortality decreased with increasing parental relative income within all birth cohorts. Among children aged 0-5, the socioeconomic gradient for all-cause, accidental deaths, sudden infant death syndrome and perinatal factors declined over the periode, while there was no systematic decline related to congenital malformations. Among children 6-21, the gradient did not weaken similarly, although there were indications of declines in the socioeconomic gradient related to all-cause deaths, accidents, and suicides. While the absolute differences in mortality declined over time, the relative differences remained stable.

**Conclusion:** There has been a large reduction in child mortality in all socioeconomic groups across 50 years for all-cause and most cause-specific mortality groups. However, children of parents in the lowest part of the income distribution still have an elevated mortality, and the relative differences have not always been declining.

## INTRODUCTION

Social differences in children’s health have been reported in a large number of studies from both rich and poor countries: most aspects of children’s health are positively linked to the parents’ socioeconomic resources, and children born into socially advantaged families have the lowest mortality (1–5). However, little is known about how the associations between childhood mortality and socioeconomic status have changed over time. The few investigations of infant mortality that have addressed this have pointed in different directions, and there is hardly any statistical evidence regarding older children (6–11). Also, it is not theoretically obvious what one should expect. On the one hand, the rising economic inequality documented in several countries including Norway (12,13) may strengthen the social gradient in mortality, depending on how it is measured. On the other hand, introduction of medical technologies that can be widely applied without regard to individual income or knowledge, such as advances in neonatology in settings with public health care, may reduce mortality and leave the social gradient unchanged or even diminished.

One problem in such research is that it is challenging to measure parental socioeconomic status consistently over time because of the secular changes in the distributions of educational and occupational attainments. Furthermore, vital statistics from many countries are not linkable to measures of individual socioeconomic status, so much current research on the topic has relied on area-level socioeconomic indicators (10,14,15).

In Norway, registers that cover the entire population include individual-level data on births, deaths, and socioeconomic resources, such as earnings. The current study aims to examine how child mortality varies with socioeconomic resources, as indicated by the parents’ ranking in the income distribution and the changes in this income gradient over a 50-year period. We use a relative income measure because of the great increase in earnings over the decades considered. In addition to all-cause mortality, 3-4 causes of death are considered. We analyse children aged 0-5 and 6-21 separately, as the causes of death and underlying behavioral and other pathways are different.

Infant mortality in Norway is lower than in most other OECD countries, and the mortality of older children is at the average (16).

## METHODS

### Study population

We used data from administrative registers covering the entire Norwegian population: the Population Register (PR) (17), the National Register for Personal Taxpayers, the Medical Birth Register (MFR) (18), and the Cause of Death Register (CDR) (19). The registers include personal identifiers that allow linkage between them, and between children and parents. Because annual income data are available from 1967 and we considered the parents’ income the year before the child was born, we included children born in Norway in 1968 or later. As the data cover the years up to 2015, the last cohort included in the analysis of mortality up to age 5 is 2010, and the last cohort included in the analysis of mortality up to 21 (conditioned on survival to age 5) is 1994. Individuals who emigrated before age 5 or 21, respectively, were excluded. Table A1 shows descriptive statistics for the study population.

### Statistical Analysis and Measures

We calculated the probability (per 1000 or 10000) of dying within the fifth birthday and, for those still alive at that time, the probability of dying within the 21^st^ birthday, by using information on birth month and year taken from the CDR or the PR. The causes of death are coded in accordance with the International Classification of Diseases, using the 8^th^, 9^th^ and 10^th^ revision. We considered four groups of causes among 0-5 year olds: perinatal factors, congenital malformations, sudden infant death syndrome and accidents, and 3 causes for children 6-21 years: accidents, suicide and cancer (see Appendix A2).

Income was measured as the sum of the mother’s and father’s reported pensionable labour income, including earnings from self-employment, in the calendar year before the child’s birth. This income was transformed into an income rank by comparing with the corresponding incomes of the parents of other children born the same year. Death probability for rank vigintiles (rank 0-5%, 5-10% etc.) were calculated for different cohort groups: 1968-79, 80-89, 90-99 and 2000-10 when considering children aged 0-5, and 1968-75, 1976-83, and 1984-94 for those aged 6-21. For each of these cohort groups we also estimated the linear relationship between parental income rank and the child death probability in a regression model, and we included pairs of cohort groups to make inferences about the changes in the linear relationship over the cohorts. A certain linear relationship is relatively stronger when the overall mortality level is high. Stated differently, the mortality of the poor divided by that of the rich may be larger in one cohort group than another even though the difference between them is the same. We define a “relative linear relationship (trend)” as the linear relationship (the slope in the regression) divided by the corresponding regression constant term (corresponding to mortality at the lowest income rank). All analyses were performed with STATA 16. (See appendix for methodological details).

## RESULTS

### All-cause Mortality Among Children Aged 0-5 Year

All-cause mortality among 0–5-year-olds declines with increasing parental relative income, as measured by vigintiles, in each of the four cohort groups (Figure 1). Table A3 shows the linear trends as well as the corresponding constant terms in the regression and the relative linear trends. There is a generally negative linear trend, and the point estimates suggest that it becomes stronger over the cohorts, although the changes over some cohort pairs are modest (see cohort interactions in Table A5). In contrast, there is a less clear pattern in the development of the relative linear trend.

**Figure 1.**
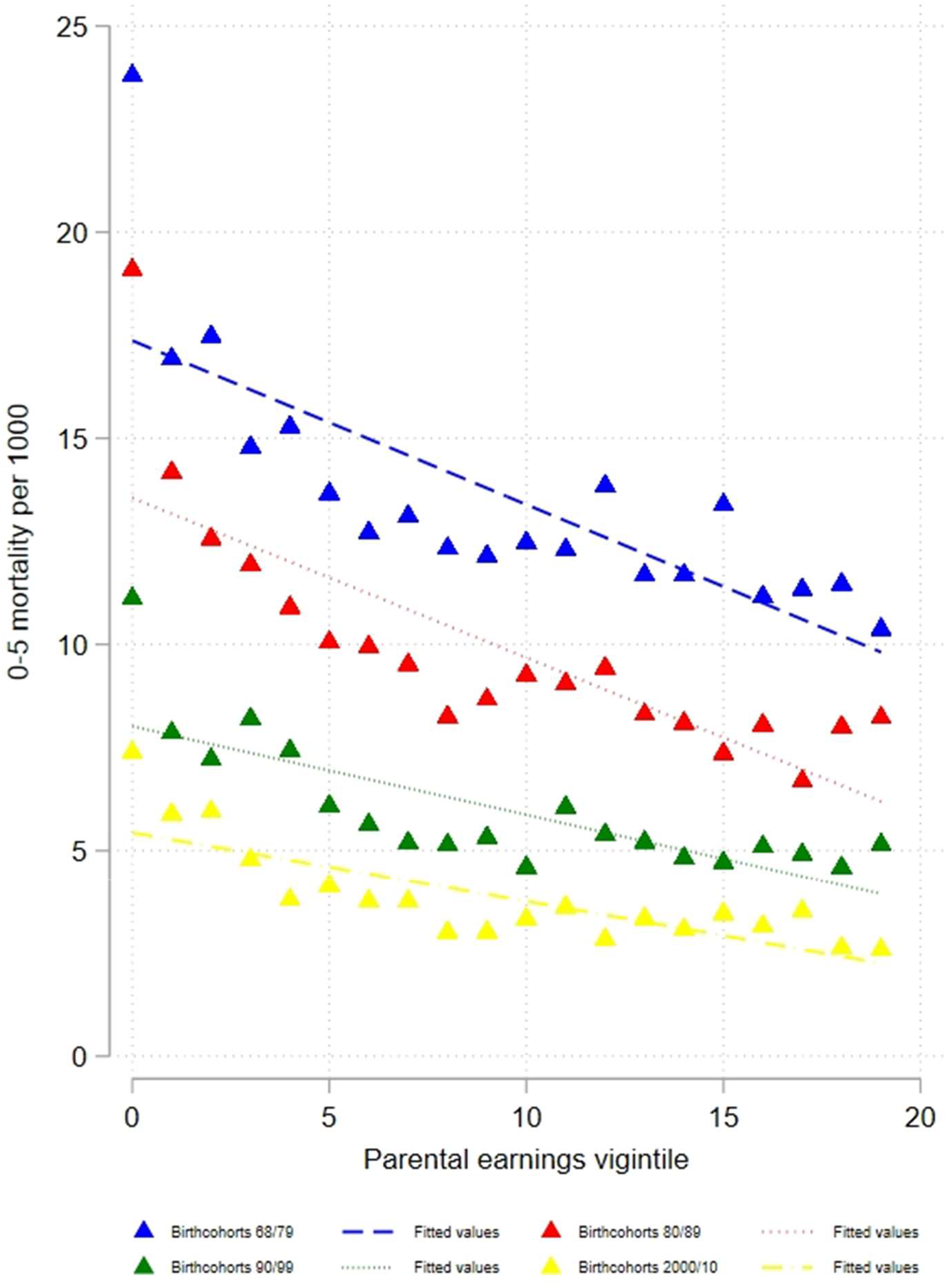
Note: Mortality rates are plotted across lowest to highest parental income vigintile (5 % groups). Straight lines refer to linear fit.

### Cause-Specific Mortality Among Children Aged 0-5 Years

As with overall mortality, the risk of dying from perinatal causes decreases with increasing parental income in all cohorts (Figure 2, panel A, and Table A3). The linear trend is significantly weaker in the 1980-1989 cohorts than in the older cohorts, but does not become further weakened in younger cohorts (although that is indicated by the point estimates). The relative trend is stable across the cohort groups. The risks of dying from congenital malformations (panel B) and SIDS also decrease with higher income, but for the former, the linear trend does not change in a systematic way over the cohorts. The linear trend becomes gradually weaker over the three youngest cohort groups for SIDS (and the relative linear trend becomes stronger). Also, the risk of accidental deaths decreases with increasing parental income in all cohort groups (panel D). This relationship becomes weaker and is hardly visible in the latest cohort group, for whom the overall level of accident mortality is very low.

**Figure 2.**
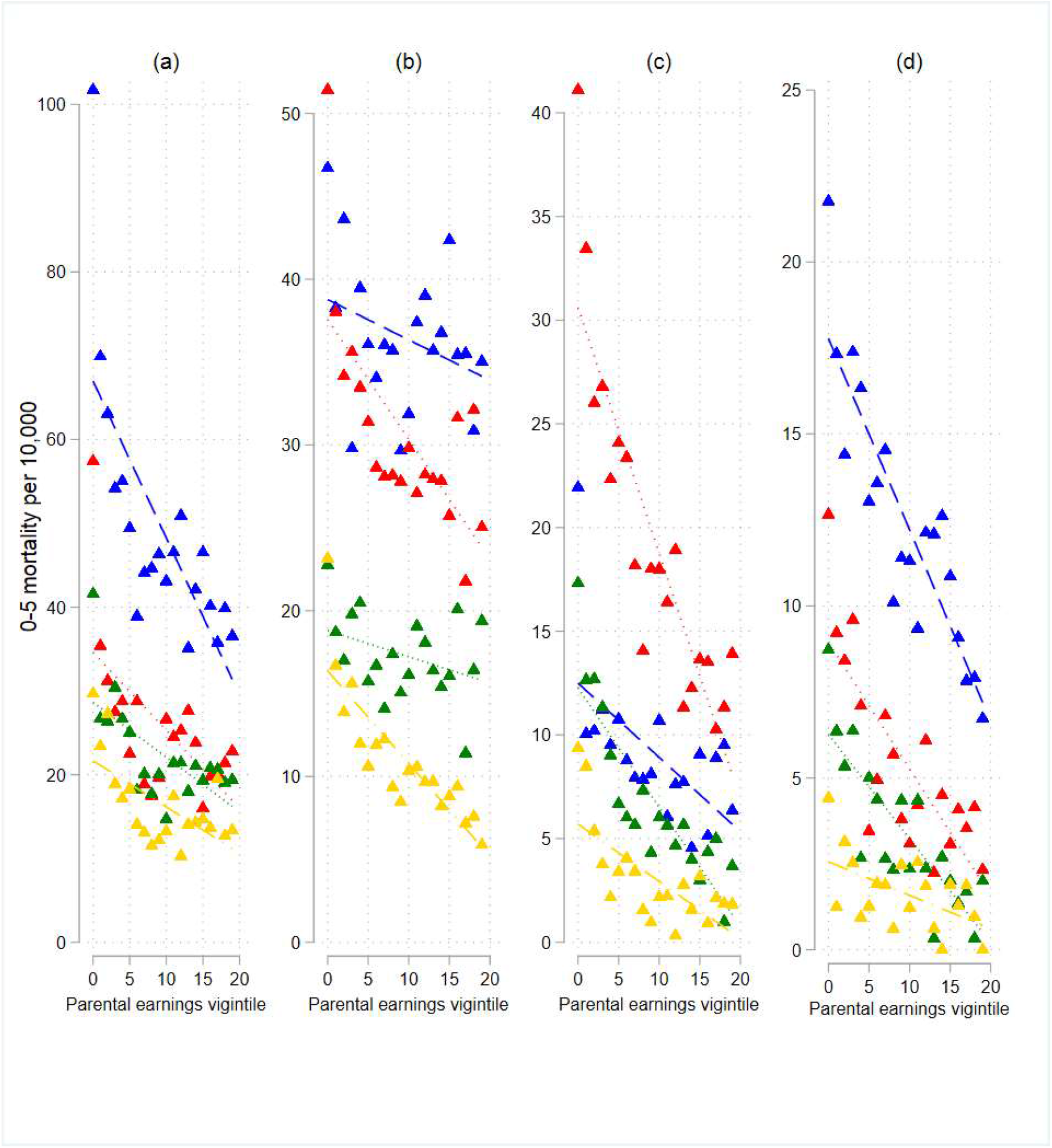
Note: Mortality rates are plotted across lowest to highest parental income vigintile (5 % groups). Blue lines:1968-1979, Red lines: 1980-1989, Green lines:1990-1999, Yellow lines: 2000-2010.

### Mortality Among Children Aged 6-21 Years

Mortality among 6–21-year-olds is inversely related to parental income in all cohort groups:1968-1975, 1976-1983 and 1984-1993 (Figure 3 and Table A4),and the point estimates of the interaction between cohort and parental income indicate that this relationship has become weaker over time. (Appendix Table A6). The relative linear trend does not change systematically.

**Figure 3.**
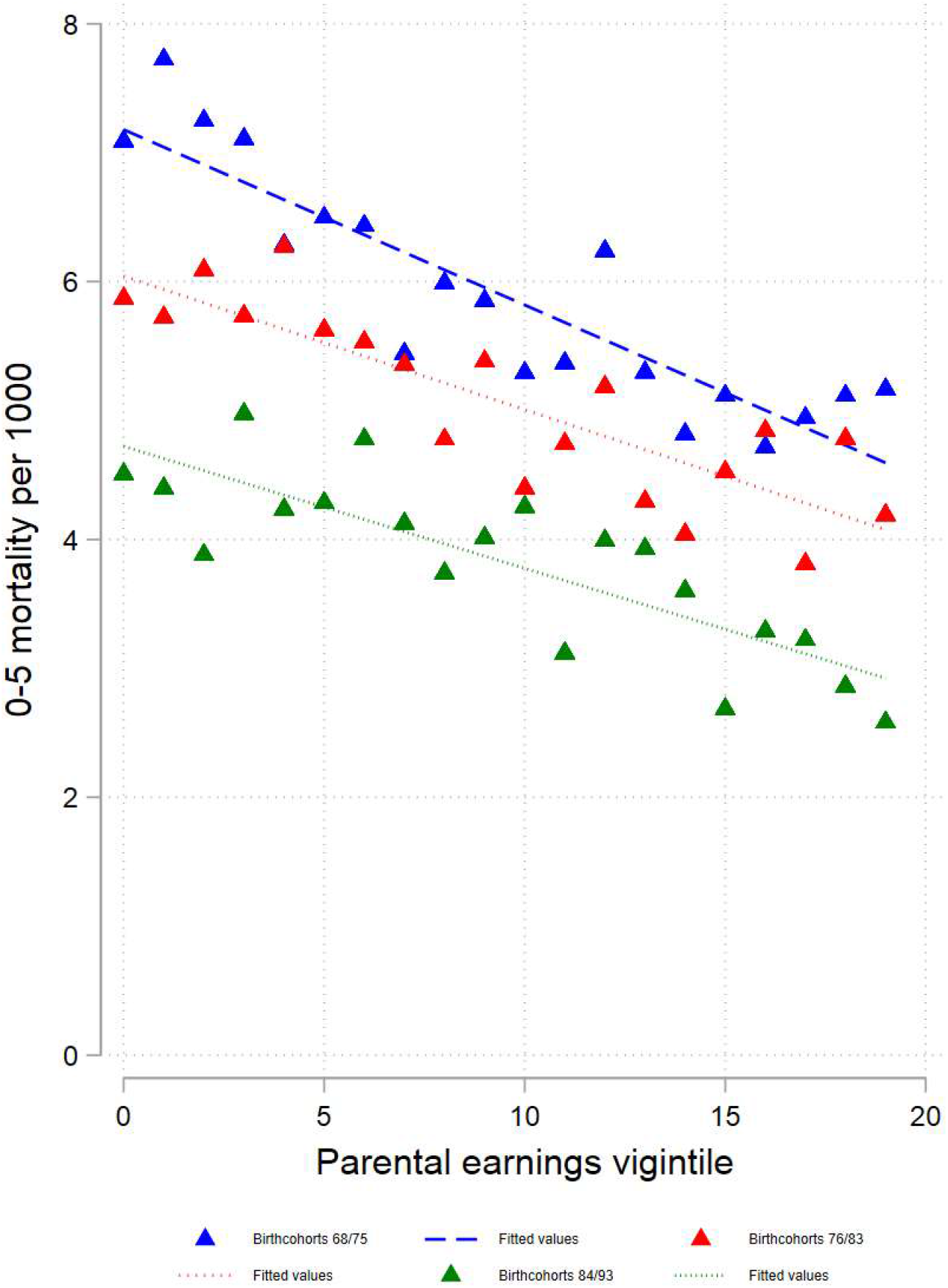
Note: Mortality rates are plotted across lowest to highest parental income vigintile (5 % groups). Straight lines refer to linear fit.

Also the mortality from accidents or suicides declines with increasing parental income. Again, the point estimates suggest a weakening relationship across the cohorts (Figure 4 and Table A4).There is a less clear development in the relative linear trends. The pattern is markedly different for cancer mortality, where an upward instead of downward gradient appears for the two oldest cohort groups.

**Figure 4.**
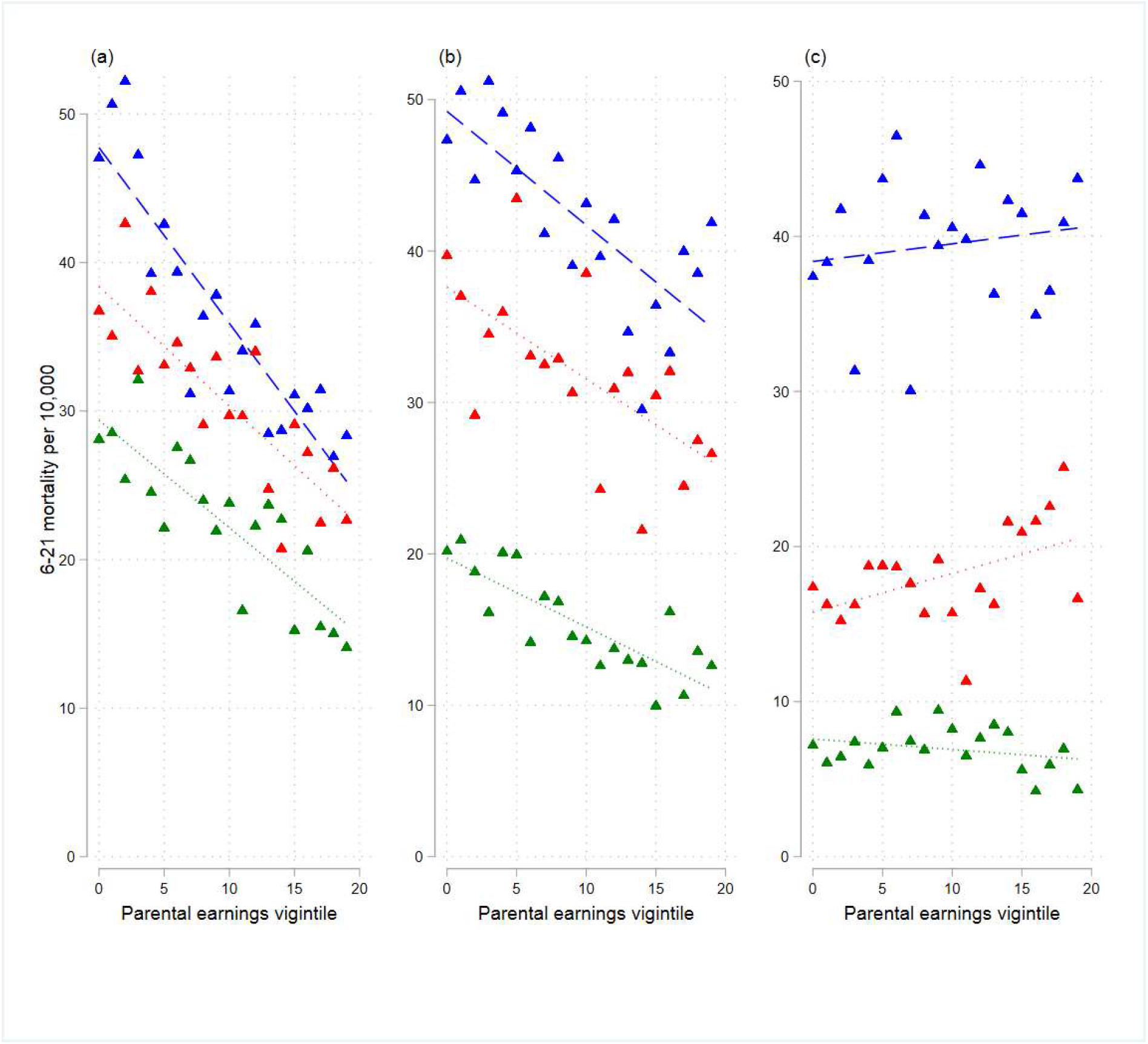
Note: Mortality rates are plotted across lowest to highest parental income vigintile (5 % groups). Straight lines refer to linear fit. Blue lines:1968-1975, Red lines: 1975-1983, Green lines:1984-1993.

## DISCUSSION

This register-based study, covering the last half-century, has shown that all-cause mortality of Norwegian children aged 0-5 or 6-21 was inversely linked to the parents’ relative income, as indicated by their position in the income distribution. The same pattern was seen in cause-of-death groups except for cancer at age 6-21.

The association between all-cause mortality and parental income probably reflects that several factors underlying childhood mortality are affected by the parents’ purchasing power or other characteristics linked to income, such as educational achievements, operating in part through, for example, knowledge of health or general analytical skills. Examples of such more proximate factors are mothers’ nutrition and smoking habits, whether the parents seek professional help when the child is sick, and whether they otherwise care well for the child. In countries without a public health care system, the association between income and use of high-quality health care may be stronger than in Norway. Additionally, an observed relationship between parents’ income and child mortality reflects that both these factors are influenced by the parents’ health in earlier years and several other individual and community characteristics.

Among children aged 0-5, the association between parents’ income and perinatal or accident mortality or (except for the oldest cohort) SIDS has become weaker, while there is no such clear trend in the mortality from congenital malformations. The income gradient in all-cause mortality (which also includes deaths from other causes than the four groups that are considered) has also become weaker. It should be noted, however, that this holds for the absolute mortality differences. The relative linear trend, which is a measure of the absolute mortality difference (trend) compared to the mortality level among the poorest, has developed in a more irregular way, except that it has become more negative for SIDS.

The probability of accident death is influenced by the parents’ protective efforts as well as various preventive public measures, such as road safety and disseminating of knowledge about home safety the (20). The general reduction of accident mortality and the weakening of the income gradient is consistent with a situation where i) parents’ protection is positively linked to income or its correlates, such as education, ii) public preventive measures are strengthened, and iii) preventive measures matter more for parents providing less protection (i.e., an interaction between these two factors).

An argument similar to that for accidents may apply for SIDS, when comparing among the three youngest cohort groups. Many countries witnessed relatively high SIDS rates in the 1980s and 1990s, which was related to advice about putting infants to sleep on their stomachs. During the early 1990s, official guidelines recommended infants instead to sleep on their back, which has been shown to be especially important for preterm and low birth weight infants typically more prevalent among low-income families (21). The information about sleep position provided to the public in the 1990s helped to eliminate this cause of death and made the earlier advantage that the well-resourced may have had (e.g., in terms of average better infant health and birth weight) gradually less relevant. However, social differences exist even in the youngest cohorts. It is likely that SIDS mortality among recent cohorts is more linked to other factors than sleeping positions, such as maternal smoking and respiratory or gastrointestinal infections (22). For this cause of death, absolute death rates were also higher among later-born cohorts than those born between 1967-1979. This likely reflects that SIDS did not have a unique ICD code until 1979, even though it was first defined in 1969 (23).

Advances in neonatal medical diagnostics and treatment have contributed to a reduced mortality from congenital malformations. The reduction has been largely the same for all income groups (as judged by the lack of a systematic weakening of the income gradient over the half century that is considered). This is as one would expect if some sub-causes were eliminated by the introduction of new medical technology and everyone had similar access to this technology.

Deaths from perinatal conditions—occurring usually within the first year of life—are largely related to low birth weight and low gestational age. The chance of premature birth is in turn influenced by, for example, maternal smoking and poor nutrition, which has typically been more common in families with low income or education (6,24). Therefore, in a hypothetical situation where smoking is almost eliminated in all population groups, mortality from perinatal conditions would go down and the income gradient would become weaker. However, this is not what has happened; smoking rates have fallen more strongly among the socioeconomically advantaged mothers (25). Also, the intake of folic acid and other vitamins is still higher in the high-income groups (26). In other words, the development in the intake of such supplements and smoking is not consistent with the change in the income gradient. Child mortality is, however, relatively rare in Norway, and it is possible that perinatal and obstetric care—provided to everyone—has increased the survival of relatively frail fetuses and children, thereby reducing the income gradient in mortality.

Among children, aged 6-21, the relationship between income and all-cause mortality has not been so clearly weakened, although there are indications in that direction. The same can be said about accident and suicide mortality, while the income gradient in the cancer mortality can be described as having become less positive and eventually negative (according to the point estimates).

As mentioned, personal risk behavior—which tends to be connected with parental socioeconomic resources (27) - is a key determinant of accident mortality (28). An example of special relevance for the relatively old children is that those whose parents have low education or income, and therefore perhaps also a relatively high tendency to live apart, may be more likely than their peers from more resourceful families to develop a drug or alcohol problem, which may lead to fatal accidents (29). The quite constant association between parental relative income and accident mortality may reflect that the links between income and the mentioned behavioural factors have been rather stable over time.

Children with low-earning parents may suffer more from mortality due to suicide because of their own or their parents’ mental health problems which in turn may be partly linked with unstable family situations (30,31). The unchanging income gradient in suicide mortality points to the persistent importance of parental income for such factors.

Socioeconomic status may be associated with certain cancer types through social patterning of risk factors such as birth weight, parental age, and environmental factors, although the direction of the association may vary (32). For example, previous research has found that the incidence of brain tumors is highest in socioeconomically advantaged residential areas, whereas some types of leukemia tend to occur more often in low socioeconomic settings (33). One reason why we see indications of a gradually weaker positive income gradient, which eventually becomes negative, may be the shift in the relative prevalence of various malignancies. While brain tumors were the most frequent diagnosis for the oldest cohort, leukemia was more prevalent for the younger ones.

### Limitations

The major strengths of our study are that i) the analysis is based on high-quality register data for an entire national population, ii) it covers several decades, iii) it involves a measure of socioeconomic resources that is particularly suitable for a study of long-term trends, and iv) it includes (unlike most earlier studies) different causes of death in addition to all-cause mortality. Finally, information on the cause of death or income is missing for less than 1% and 0.2 % of the sample, respectively.

Changes in coding practice constitute a limitation because they make it difficult to understand the development with respect to SIDS. Furthermore, children born abroad are not included since we do not know the parents’ income the year before birth. Perhaps more importantly, the chance of losing a child in a certain year is influenced by the parents’ resources at that time and some years back, which may not be adequately captured by the income a year before birth, when the parents may, for example, have earned little because of studies. That said, it would not have been a good idea to consider parental income at, say, age five instead, as this might have been affected by an earlier child death (i.e. reverse causality).

Parental education is also known to be an important correlate of children’s mortality (34,35), but the strong educational expansion over time calls for a relative measure, and this would be difficult to construct because education is not a continuous variable such as income; only a few educational categories are defined. Another potential problem with the present study is that the parents may not live together. In that case, the resources from which the child benefits may be smaller than suggested by the sum of the mother’s and father’s income. One possible argument is that the increasing prevalence of union dissolution would make our income measure an increasingly poor indicator of the income rank in later years. In other words, the importance of the current purchasing power for child mortality would – especially for the youngest cohorts – be larger than indicated by our results based on the pre-birth income.

Finally, the relationship between income and child mortality is not necessarily linear, even though a linear trend was calculated for simplicity. On the contrary, the graphs suggest, as one would expect, that increasing income matters less at the higher levels.

An implication of the study is that there have been large reductions in the social gradient in child mortality over the last half century. However, children whose parents are in the lowest part of the income distribution still have elevated mortality, and the income gradient has not been declining for all causes of death that have been considered. Thus, despite the overall progress, a disadvantage associated with low income, seems difficult to overcome.

## Data Availability

Data are not publicly available; but can be accessed through Statistics Norway and the Norwegian Institute for Public Health by permission upon relevant approvals.

## Research Ethics Approval

The study has been approved by the Norwegian Regional Committees for Medical and Health Research Ethics (REC), approval number 2013/2394.

## Key points

- Child mortality has declined over the last 50 years, but few studies have examined if the decline has been equal across socioeconomic groups.
- We addressed this issue by using a relative measure of parental income before birth.
- The absolute income inequality in child mortality has decreased for all-cause mortality and some causes of death, but more clearly at age 0-5 than at age 6-21.
- The *relative* difference in mortality between income groups has been more stable.

## REFERENCES

1. Case A, Lubotsky D, Paxson C. Economic Status and Health in Childhood: The Origins of the Gradient. Am Econ Rev. 2002 Dec;92(5):1308–34.

2. Gupta RP-S, de Wit ML, McKeown D. The impact of poverty on the current and future health status of children. Paediatr Child Health. 2007 Oct;12(8):667–72.

3. Grøholt E-K, Stigum H, Nordhagen R, Köhler L. Children with chronic health conditions in the Nordic countries in 1996 – influence of socio-economic factors. Ambul Child Health. 2001;7(3–4):177–89.

4. Finch BK. Early Origins of the Gradient: The Relationship between Socioeconomic Status and Infant Mortality in the United States. Demography. 2003;40(4):675–99.

5. Dowd JB, Zajacova A, Aiello A. Early origins of health disparities: burden of infection, health, and socioeconomic status in U.S. children. Soc Sci Med 1982. 2009 Feb;68(4):699–707.

6. Nath S, Hardelid P, Zylbersztejn A. Are infant mortality rates increasing in England? The effect of extreme prematurity and early neonatal deaths. J Public Health [Internet]. 2020 Mar 2 [cited 2020 Nov 13]; Available from: https://academic.oup.com/jpubhealth/advance-article/doi/10.1093/pubmed/fdaa025/5771308

7. Lewer D, Jayatunga W, Aldridge RW, Edge C, Marmot M, Story A, et al. Premature mortality attributable to socioeconomic inequality in England between 2003 and 2018: an observational study. Lancet Public Health. 2020 Jan 1;5(1):e33–41.

8. Taylor-Robinson D, Lai ETC, Wickham S, Rose T, Norman P, Bambra C, et al. Assessing the impact of rising child poverty on the unprecedented rise in infant mortality in England, 2000–2017: time trend analysis. BMJ Open. 2019 Oct 1;9(10):e029424.

9. Butikofer A, Karadakic R, Salvanes KG. kIncome Inequality and Mortality: A Norwegian Perspective [Internet]. Rochester, NY: Social Science Research Network; 2021 Jan [cited 2021 Mar 6]. Report No.: ID 3774001. Available from: https://papers.ssrn.com/abstract=3774001

10. Currie J, Schwandt H. Inequality in mortality decreased among the young while increasing for older adults, 1990–2010. Science. 2016;352(6286):708–12.

11. Nair H, Byass P. Mortality in older children and adolescents: the forgotten ones. Lancet Child Adolesc Health. 2018 May;2(5):306–7.

12. In It Together: Why Less Inequality Benefits All | en | OECD [Internet]. [cited 2021 Apr 19]. Available from: https://www.oecd.org/social/in-it-together-why-less-inequality-bene-fits-all-9789264235120-en.htm

13. Markussen S, Røed K. Economic Mobility Under Pressure. J Eur Econ Assoc [Internet]. 2019 Sep 10 [cited 2019 Dec 2]; Available from: https://academic.oup.com/jeea/advance-article/doi/10.1093/jeea/jvz044/5567255

14. Singh GK, Kogan MD. Widening Socioeconomic Disparities in US Childhood Mortality, 1969–2000. Am J Public Health. 2007;Sep;97(9):1658–65.

15. Colvin JD, Zaniletti I, Fieldston ES, Gottlieb LM, Raphael JL, Hall M, et al. Socioeconomic Status and In-Hospital Pediatric Mortality. Pediatrics. 2013 Jan 1;131(1):e182–90.

16. Thakrar AP, Forrest AD, Maltenfort MG, Forrest CB. Child Mortality In The US And 19 OECD Comparator Nations: A 50-Year Time-Trend Analysis. Health Aff (Millwood). 2018 Jan;37(1):140–9.

17. Dette er Folkeregisteret [Internet]. Skatteetaten. [cited 2021 Nov 29]. Available from: https://www.skatteetaten.no/person/folkeregister/om/om/

18. Medisinsk fødselsregister [Internet]. Folkehelseinstituttet. [cited 2021 Nov 29]. Available from: https://www.fhi.no/hn/helseregistre-og-registre/mfr/

19. Dødsårsaksregisteret [Internet]. Folkehelseinstituttet. [cited 2021 Nov 29]. Available from: https://www.fhi.no/hn/helseregistre-og-registre/dodsarsaksregisteret/

20. Glied SA. The Value of Reductions in Child Injury Mortality in the United States. In: Medical Care Output and Productivity [Internet]. University of Chicago Press; 2001 [cited 2021 Nov 7]. p. 511–38. Available from: https://www.nber.org/books-and-chapters/medi-cal-care-output-and-productivity/value-reductions-child-injury-mortality-united-states

21. Altindag O, Greve J, Tekin E. Public Health Policy At Scale: Impact of a Government-sponsored Information Campaign on Infant Mortality in Denmark [Internet]. National Bureau of Economic Research; 2021 Mar [cited 2021 Dec 2]. (Working Paper Series). Report No.: 28621. Available from: https://www.nber.org/papers/w28621

22. Moon RY, Hauck FR. SIDS Risk: It’s More Than Just the Sleep Environment. Pediatrics [Internet]. 2016 Jan 1 [cited 2021 Nov 5];137(1). Available from: https://pediatrics.aap-publications.org/content/137/1/e20153665

23. Shapiro-Mendoza CK, Parks S, Lambert AE, Camperlengo L, Cottengim C, Olson C. The Epidemiology of Sudden Infant Death Syndrome and Sudden Unexpected Infant Deaths: Diagnostic Shift and other Temporal Changes. In: Duncan JR, Byard RW, editors. SIDS Sudden Infant and Early Childhood Death: The Past, the Present and the Future [Internet]. Adelaide (AU): University of Adelaide Press; 2018 [cited 2021 Sep 17]. Available from: http://www.ncbi.nlm.nih.gov/books/NBK513373/

24. Kramer MS. Determinants of low birth weight: methodological assessment and meta-analysis. Bull World Health Organ. 1987;65(5):663–737.

25. Grøtvedt L, Kvalvik LG, Grøholt E-K, Akerkar R, Egeland GM. Development of social and demographic differences in maternal smoking between 1999 and 2014 in Norway. Nicotine Tob Res. 2017;19(5):539–46.

26. Nilsen RM, Vollset SE, Gjessing HK, Magnus P, Meltzer HM, Haugen M, et al. Patterns and predictors of folic acid supplement use among pregnant women: the Norwegian Mother and Child Cohort Study. Am J Clin Nutr. 2006 Nov 1;84(5):1134–41.

27. Kipping RR, Smith M, Heron J, Hickman M, Campbell R. Multiple risk behaviour in adolescence and socio-economic status: findings from a UK birth cohort. Eur J Public Health. 2015 Feb 1;25(1):44–9.

28. Tarkiainen L, Martikainen P, Laaksonen M, Aaltonen M. Childhood family background and mortality differences by income in adulthood: fixed-effects analysis of Finnish siblings. Eur J Public Health. 2015 Apr;25(2):305–10.

29. Martikainen P, Elo I, Tarkiainen L, Mikkonen J, Myrskylä M, Moustgaard H. The changing contribution of childhood social characteristics to mortality: a comparison of Finnish cohorts born in 1936–50 and 1961–75. Int J Epidemiol [Internet]. [cited 2020 May 6]; Available from: https://academic.oup.com/ije/advance-arti-cle/doi/10.1093/ije/dyaa041/5816047

30. Evans GW. The environment of childhood poverty. Am Psychol. 2004;59(2):77.

31. the STOP Consortium, Carballo JJ, Llorente C, Kehrmann L, Flamarique I, Zuddas A, et al. Psychosocial risk factors for suicidality in children and adolescents. Eur Child Adolesc Psychiatry. 2020 Jun;29(6):759–76.

32. Spector LG, Pankratz N, Marcotte EL. Genetic and nongenetic risk factors for childhood cancer. Pediatr Clin North Am. 2015 Feb;62(1):11–25.

33. Alston RD, Rowan S, Eden TOB, Moran A, Birch JM. Cancer incidence patterns by region and socioeconomic deprivation in teenagers and young adults in England. Br J Cancer. 2007 Jun;96(11):1760–6.

34. Arntzen A. Socioeconomic status and risk of infant death. A population-based study of trends in Norway, 1967-1998. Int J Epidemiol. 2004 Apr 1;33(2):279–88.

35. Rom AL, Mortensen LH, Cnattingius S, Arntzen A, Gissler M, Andersen A-MN. A comparative study of educational inequality in the risk of stillbirth in Denmark, Finland, Norway and Sweden 1981–2000. J Epidemiol Community Health. 2012;66(3):240–6.

