## Supplementary data for "Parental income gradients in child and adolescent mortality: Norwegian trends over half a century"

This file includes details on coding of causes of death, estimates for cohort changes and a specification of the linear probability models that are estimated.

**Supplementary tables:**

Table A1: Descriptive statistics for children born in Norway 1968-2010

Table A2: Selected causes of deaths and codes

Table A3: All cause and cause-specific estimates for mortality among 0-5 yrs old

Table A4: All cause and cause-specific estimates for mortality among 6-21 yrs old

Table A5: Estimated differences in the linear association between all-cause and cause-specific child mortality, ages 0-5, and parental income vigintile by birth cohort period.

Table A6: Estimated differences in the linear association between all-cause and cause-specific child mortality, ages 6-21, and parental income vigintile by birth cohort period.

Table A1: Descriptive statistics for children born in Norway 1968-2010

Panel A. Deaths 0-5 years

| | Birth cohort 1969-79 | Birth cohort 1980-1989 | Birth cohort 1990-1999 | Birth cohort 2000-2010 |
|---|---|---|---|---|
| Total number of deaths | 9.866 | 5190 | 3578 | 2460 |
| Percent | 1.38 | 0.99 | 0.60 | 0.38 |
| Parental income (mean) | 307 521.2 | 413705.4 | 485911.8 | 673848.8 |
| St.dev | 156426.1 | 204624.1 | 302092.7 | 410707.5 |
| Individuals | 713.999 | 525.869 | 597.394 | 641.51 |

Panel B. Deaths 6-21 years

| | Birth cohort 1968-1975 | Birth cohort 1976-1983 | Birth cohort 1984-1993 |
|---|---|---|---|
| Total number of deaths | 2.953 | 2.050 | 2.122 |
| Percent | 0.59 | 0.51 | 0.38 |
| Parental income (mean) | 283037.3 | 379384.2 | 438405.8 |
| St.dev | 143176 | 178484 | 227452.3 |
| Individuals | 506.574 | 405.327 | 564.449 |

Table A1: Selected causes of death and codes

| | ICD-101 (1996-) | ICD-9 (1986-95) | ICD-8 (1969-85) |
|---|---|---|---|
| Sudden infant death syndrome | R95 | 798.0 | 795 and age |
| External causes of injury and poisoning | V01-Y89 | E800-E999 | E800-E999 |
| Suicide | X60-X84 | E950-E959 | E950-E959 |
| Cancer | C00-C97 | 140-208 | 140-209 |
| Certain conditions originating in the perinatal period | P00-P96 | 760-779 | 760-779 |
| Congenital malformations and chromosomal abnormalities | Q00-Q99 | 740-759 | 740-759 7 |

Table A3: All cause and cause-specific estimates for mortality among 0-5 yrs old

| | Constant | Slope | Lower 95% CI | Upper 95% CI | Relativ slope |
|---|---|---|---|---|---|
| (a) All-cause | (1) | (2) | (3) | (4) | (5) |
| 1968-1979 | 0.0176208 | **-0.0004003** | -0.0004472 | -0.0003533 | -0.022717470 |
| 1980-1989 | 0.0135607 | **-0.0003886** | -0.0004349 | -0.0003422 | -0.028656338 |
| 1990-1999 | 0.0080268 | **-0.0002145** | -0.0002484 | -0.0001805 | -0.026722978 |
| 2000-2010 | 0.0054198 | **-0.0001669** | -0.0001931 | -0.0001406 | -0.030794494 |
| (b) Prental conditions | | | | | |
| 1968-1979 | 0.0068216 | **-0.0001912** | -0.0002195 | -0.0001628 | -0.028028615 |
| 1980-1989 | 0.0034734 | **-0.0000930** | -0.0001168 | -0.0000692 | -0.026774918 |
| 1990-1999 | 0.0028658 | **-0.0000652** | -0.0000860 | -0.0000444 | -0.022751064 |
| 2000-2010 | 0.0021637 | **-0.0000550** | -0.0000721 | -0.0000378 | -0.02541942 |
| (c) Congenital malformations | | | | | |
| 1968-1979 | 0.0038560 | **-0.0000227** | -0.000047 | 0.0000015 | -0.005886929 |
| 1980-1989 | 0.0037283 | **-0.0000718** | -0.0000976 | -0.0000459 | -0.019258107 |
| 1990-1999 | 0.0018820 | **-0.0000159** | -0.0000342 | 0.0000024 | -0.008448459 |
| 2000-2010 | 0.0016328 | **-0.0000559** | -0.0000699 | -0.0000418 | -0.034235669 |
| (d) Sudden infant death syndrome | | | | | |
| 1968-1979 | 0.0012093 | **-0.0000349** | -0.0000468 | -0.0000229 | -0.028859671 |
| 1980-1989 | 0.0030866 | **-0.0001199** | -0.0001406 | -0.0000993 | -0.038845331 |
| 1990-1999 | 0.0012315 | **-0.0000577** | -0.0000692 | -0.0000463 | -0.046853431 |
| 2000-2010 | 0.0005674 | **-0.0000274** | -0.0000348 | -0.000020 | -0.048290448 |
| (e) Accidents | | | | | |
| 1968-1979 | 0.0018235 | **-0.0000568** | -0.0000712 | -0.0000424 | -0.031148889 |
| 1980-1989 | 0.0008910 | **-0.0000365** | -0.0000475 | -0.0000256 | -0.040965208 |
| 1990-1999 | 0.0006270 | **-0.0000304** | -0.0000385 | -0.0000223 | -0.048484848 |
| 2000-2010 | 0.0002539 | **-0.0000097** | -0.0000151 | -0.0000043 | -0.038046475 |

Note: columns 1 and 2 show the constant and slope (linear trend) when mortality is regressed on parental income rank with a linear probability model. Lower 95 per cent CI is given i column 3 and upper in column 4. Column 5 shows the relative slope defined as slope divided by constant. Coeffcients in bold are significant at $P<0.05$ level.

Table A4:All-cause and cause-specific estimates for mortality among 6-21 year olds

| | Constant | Slope | Lower 95% CI | Upper 95% CI | Relative slope |
|---|---|---|---|---|---|
| (a) All-cause | (1) | (2) | (3) | (4) | (5) |
| 1968-1975 | 0.0072115 | **-0.0001360** | -0.0001729 | -0.0000990 | -0.018858767 |
| 1976-1983 | 0.0060453 | **-0.0001038** | -0.0001417 | -0.0000660 | -0.017170364 |
| 1984-1993 | 0.0047099 | **-0.0000965** | -0.0001244 | -0.0000685 | -0.020488758 |
| (b)External cause | | | | | |
| 1968-1975 | 0.0036691 | **-0.0000815** | -0.0001073 | -0.0000556 | -0.022212532 |
| 1976-1983 | 0.0027284 | **-0.0000594** | -0.0000842 | -0.0000346 | -0.021771001 |
| 1984-1993 | 0.0020319 | **-0.0000509** | -0.0000688 | -0.0000330 | -0.025050445 |
| (c)Suicide | | | | | |
| 1968-1975 | 0.0049323 | **-0.0000746** | -0.0001058 | -0.0000434 | -0.01512479 |
| 1976-1983 | 0.0037658 | **-0.0000605** | -0.0000907 | -0.0000304 | -0.016065643 |
| 1984-1993 | 0.0019310 | -0.0000446 | -0.0000623 | -0.0000270 | -0.023096841 |
| (d) Cancer | | | | | |
| 1968-1975 | 0.0038820 | 0.0000116 | -0.0000188 | 0.0000419 | 0.00298815 |
| 1976-1983 | 0.0015828 | **0.0000250** | 0.0000022 | 0.0000478 | 0.015794794 |
| 1984-1993 | 0.0007525 | -0.0000073 | -0.0000192 | 0.0000045 | -0.009754153 |

Note: columns 1 and 2 show the constant and slope (linear trend) when mortality is regressed on parental income rank with a linear probability model. Lower 95 per cent CI is given i column 3 and upper in column 4. Column 5 shows the relative slope defined as slope divided by constant. Coeffcients in bold are significant at P<0.05 level.

**Statistical models**

For all cause and cause-specific measures of child mortality, we estimate the following linear regression model:

$$M_i = \beta_0 + \beta_1 \text{Parental income}_i + \varepsilon_i \quad (1)$$

where $M_i$ is 1 if child i dies from the mortality outcome under study within the relevant observation period (age 0-5 or age 6-21) and otherwise is 0. *Parental income*$_i$ is the parental income rank, which is a number between 0 (the poorest parents) to 1 (the richest parents). The coefficient of interest is $\beta_1$, which is referred to as the main text as the income gradient, or the linear relationship (trend) between relative income and mortality.

Estimates of $\beta_1$ and the corresponding 95% confidence intervals are shown in tables along with the relative linear relationships (relative slopes) defined as $\beta_1$ divided by $\beta_0$.

To find out whether the income gradient changes over cohorts, an interaction between Parental income and cohort group was added along with the cohort main effect. More specifically, the following model was estimated for all cohorts combined:

$$M_i = \beta_0 + \beta_1 \text{Parental income}_i + \beta_2 \text{Parental income}_i * K_i + \beta_3 K_i \, \varepsilon_i \quad (2)$$

$K_i$ is a categorical birth cohort variable with the oldest cohort group as the reference (i.e. when analysing mortality between ages 0 and 5, the birth cohort 1968-1979 serves as the reference group, and when the focus is on mortality between ages 6 and 21, 1968-1975 serves as the reference category).

The $\beta_2$ coefficients are shown in tables A5 and A6.

Table A5: Estimated differences in the linear association between all-cause and cause-specific child mortality, ages 0-5, and parental income vigintile by birth period.

| | (a) All-cause mortality | | | (b) Perinatal | | | (c) Congenital | | | (d) SIDS | | | (e) Accidents | | |
|---|---|---|---|---|---|---|---|---|---|---|---|---|---|---|---|
| | Coef. | Lower 95% CI | Upper 95% CI | Coef. | Lower 95% CI | Upper 95% CI | Coef. | Lower 95% CI | Upper 95% CI | Coef. | Lower 95% CI | Upper 95% CI | Coef. | Lower 95% CI | Upper 95% CI |
| Vigintile (linear) | -0.00040 | -0.00044 | -0.00036 | -0.00019 | -0.00021 | -0.00017 | -0.00002 | -0.00004 | 0.00000 | -0.00003 | -0.00005 | -0.00002 | -0.00006 | -0.00007 | -0.00005 |
| Cohorts (ref. = 1968-1979) | | | | | | | | | | | | | | | |
| Cohort 1980-1989 | -0.00406 | -0.00469 | -0.00343 | -0.00335 | -0.00372 | -0.00298 | -0.00013 | -0.00046 | 0.00021 | 0.00188 | 0.00167 | 0.00208 | -0.00093 | -0.00110 | -0.00076 |
| Cohort 1990-1999 | -0.00959 | -0.01020 | -0.00899 | -0.00396 | -0.00432 | -0.00360 | -0.00197 | -0.00230 | -0.00165 | 0.00002 | -0.00018 | 0.00022 | -0.00120 | -0.00136 | -0.00103 |
| Cohort 2000-2010 | -0.01220 | -0.01280 | -0.01160 | -0.00466 | -0.00501 | -0.00431 | -0.00222 | -0.00254 | -0.00191 | -0.00064 | -0.00084 | -0.00045 | -0.00157 | -0.00173 | -0.00141 |
| Interaction terms | | | | | | | | | | | | | | | |
| Vigintile x Cohort 1980-1989 | 0.00001 | -0.00004 | 0.00007 | **0.00010** | 0.00006 | 0.00013 | **-0.00005** | -0.00008 | -0.00002 | **-0.00009** | -0.00010 | -0.00007 | **0.00002** | 0.00000 | 0.00004 |
| Vigintile x Cohort 1990-1999 | **0.00019** | 0.00013 | 0.00024 | **0.00013** | 0.00009 | 0.00016 | 0.00001 | -0.00002 | 0.00004 | **-0.00002** | -0.00004 | 0.00000 | **0.00003** | 0.00001 | 0.00004 |
| Vigintile x Cohort 2000-2010 | **0.00023** | 0.00018 | 0.00029 | **0.00014** | 0.00010 | 0.00017 | **-0.00003** | -0.00006 | 0.00000 | 0.00001 | -0.00001 | 0.00003 | **0.00005** | 0.00003 | 0.00006 |
| Intercept | 0.01762 | 0.01721 | 0.01803 | 0.00682 | 0.00658 | 0.00706 | 0.00386 | 0.00364 | 0.00407 | 0.00121 | 0.00107 | 0.00134 | 0.00182 | 0.00171 | 0.00193 |
| N | 2 478 772 | | | 2 478 772 | | | 2 478 772 | | | 2 478 772 | | | 2 478 772 | | |

Note: Coefficients in bold are significant athe *P* <0.05-level.

Table A6: Estimated differences in the linear association between all-cause and cause-specific child mortality, ages 6-21, and parental income vigintile by birth period.

| | (a) All-cause mortality | | | (b) External cause | | | (c) Suicides | | | (d) Cancer | | |
|---|---|---|---|---|---|---|---|---|---|---|---|---|
| | Coef. | Lower 95% CI | Upper 95% CI | Coef. | Lower 95% CI | Upper 95% CI | Coef. | Lower 95% CI | Upper 95% CI | Coef. | Lower 95% CI | Upper 95% CI |
| Vigintile (linear) | -0.00014 | -0.00017 | -0.00010 | 0.00001 | -0.00008 | -0.00003 | -0.00006 | -0.00009 | -0.00003 | 0.00003 | 0.00000 | 0.00005 |
| Cohorts (ref. = 1968-1975) | | | | | | | | | | | | |
| Cohort 1976-1983 | -0.00117 | -0.00172 | -0.00061 | 0.00094 | 0.00057 | 0.00131 | 0.00117 | 0.00074 | 0.00160 | 0.00230 | 0.00193 | 0.00267 |
| Cohort 1984-1993 | -0.00250 | -0.00301 | -0.00199 | -0.00070 | -0.00106 | -0.00033 | -0.00183 | -0.00225 | -0.00142 | -0.00083 | -0.00119 | -0.00047 |
| Interaction terms | | | | | | | | | | | | |
| Vigintile x Cohort 1976-1983 | 0.00003 | -0.00002 | 0.00008 | -0.00002 | -0.00006 | 0.00001 | -0.00001 | -0.00005 | 0.00002 | -0.00001 | -0.00005 | 0.00002 |
| Vigintile x Cohort 1984-1993 | 0.00004 | -0.00001 | 0.00009 | 0.00001 | -0.00002 | 0.00004 | 0.00002 | -0.00002 | 0.00005 | **-0.00003** | -0.00006 | 0.00000 |
| Intercept | 0.00721 | 0.00684 | 0.00758 | 0.00273 | 0.00245 | 0.00301 | 0.00377 | 0.00345 | 0.00409 | 0.00158 | 0.00131 | 0.00186 |
| N | 1 463 869 | | | 1 463 869 | | | 1 463 869 | | | 1 463 869 | | |

Note: Coefficients in bold are significant athe *P* <0.05-level.
